# Humoral response to Pfizer mRNA vaccine against SARS CoV2, in patients with autoimmune inflammatory rheumatic diseases and the impact on the rheumatic disease activity

**DOI:** 10.1101/2021.04.02.21254493

**Authors:** Yolanda Braun-Moscovici, Marielle Kaplan, Doron Markovits, Samy Giryes, Kochava Toledano, Yonit Tavor, Katya Dolnikov, Alexandra Balbir-Gurman

## Abstract

**Background:** The registration trials of mRNA vaccines against SARS CoV2 did not address patients with autoimmune inflammatory rheumatoid diseases (AIRD).

**Aims:** To assess the humoral response to mRNA vaccine against SARS CoV2, in AIRD patients treated with immunomodulating drugs and the impact on AIRD activity.

**Methods:** Consecutive patients treated at the rheumatology institute who received their first SARS-CoV-2 (Pfizer) vaccine were recruited to the study, at their routine visit. The patients were invited for serology test 4-6 weeks after receiving the second dose of vaccine. IgG Antibodies (Ab) against SARS COV2 virus were detected using the SARS-Cov-2 IgG II Quant (Abbott) assay

**Results:** One hundred fifty-six consecutive patients (76% females) treated at a single rheumatology center (mean age (range) 59.1 (21-83) years), mean (range) disease duration 10.8 (1-55) years), were recruited to the study. Thirty-five percents of patients received conventional synthetic (cs)DMARDs only, 64% biological/targeted synthetic (b/ts) DMARDs, 34% received combined treatment with csDMARDs and b/tsDMARDs and 32% corticosteroids (mean dose(range) 5.8mg(2.5-20mg) prednisone). One hundred thirty-seven patients (88%) were seropositive for IgG Ab against SARS CoV2 virus (median 2832.5 AU/ml, range 58-29499). Nineteen (12%) patients had negative tests, 11/19 were treated with B cell depleting agents. The reported side effects of the vaccine were minor (muscle sore, headache, low grade fever). The rheumatic disease remained stable in all patients.

**Conclusions:** The vast majority of AIRD patients developed a significant humoral response following the administration of the second dose of the Pfeizer mRNA vaccine against SARS CoV2 virus. Only minor side effects were reported and no apparent impact on AIRD activity was noted.

---

The registration trials of mRNA vaccines against SARS CoV2 did not address patients with autoimmune inflammatory rheumatic diseases (AIIRD)(1,2). Concerns were raised whether these patients can mount a protective immune response and whether the vaccination may trigger a flare up of the AIIRD. Previous studies showed that most protein based vaccines induce significant humoral responses in AIIRD patients that reach protective antibody titers (3). However, the humoral response was found to be blunted in some of the patients treated with immunosuppressive therapy or with CD20-depleting antibodies (4). We wish to report the humoral response to mRNA vaccine against SARS CoV2, in AIIRD patients treated with Disease Modifying Anti Rheumatic Drugs (DMARDs) and the impact on AIIRD activity.

One hundred fifty-six consecutive patients (76% females) treated at a single rheumatology center (mean age (range) 59.1 (21-83) years), mean (range) disease duration 10.8 (1-55) years), who received their first SARS-CoV-2 (Pfizer) vaccine were recruited to the study, at their routine visit. The visit included AIIRD activity assessment and questioning regarding vaccine side effects. The patients were invited for serology tests 4-6 weeks after the second dose of vaccine. IgG Antibodies (Ab) against SARS COV2 virus were detected using the SARS-Cov-2 IgG II Quant (Abbott) assay based on a chemiluminescent microparticle immunoassay (CMIA) on the ARCHITECT ci8200system from Abbott. This assay measures IgG antibodies against the spike receptor-binding domain (RBD) of the virus; the test is considered positive above 50 AU/ml. The study was approved by the local ethical committee.

The cohort included 78 patients with inflammatory joint disease (rheumatoid arthritis-47, psoriatic arthropathy-19, spondyloarthopathy-9, juvenile rheumatoid arthritis-2, sarcoidosis-1), 18 patients with systemic lupus erythematosus (5 of them had also antiphospholipid syndrome), 46 patients with other connective tissue diseases (systemic sclerosis-36, inflammatory myopathy-9, Sjogren disease-2, mixed connective tissue disease-1), 10 patients with vasculitis (Takayasu’s disease-4, granulomatosis with polyangiitis-2, eosinophilic granulomatous polyangiitis-2, Behcet’s syndrome-2), IgG4 related disease and idiopathic recurrent pericarditis (1 patient each). thirty-five percents of patients received conventional synthetic (cs)DMARDs only, 64% biological/targeted synthetic (b/ts) DMARDs, 34% received combined treatment with csDMARDs and b/tsDMARDs and 32% corticosteroids (mean dose(range) 5.8mg(2.5-20mg) prednisone). One hundred thirty-seven patients (88%) were seropositive for IgG Ab against SARS CoV2 virus (median 2832.5 AU/ml, range 58-29499). Nineteen (12%) patients had negative tests: 9 out of 19 rituximab treated patients (1.5-12 months before), 2 out of 2 abatacept treated patients, 3 out of 15 patients treated with mycophenolate mofetil (MMF) only, 1 out of 6 belimumab treated patients (the patient also received MMF), 1 patient treated with secukinumab, 1 patient treated with prednisone 20 mg, 1 patient treated with chemotherapy for a lung neoplasm and the only patient treated with obinutuzumab. The antibody titers correlated with the type of immunomodulatory treatment and not with the AIIRD diagnosis or the patients age (Figure1). The IgG Ab titers were higher in the vaccinated patients compared to 15 recovering COVID 19 AIIRD patients (mean (median) 4956.2(2221)AU/ml vs 799.2(211)AU/ml (Figure2).The reported side effects of the vaccine were minor (muscle sore, headache, low grade fever). One patient with Familial Mediterranean Fever, interstitial lung disease and positive rheumatoid factor reported of new onset arthritis 2 weeks after the first dose of vaccine. No flare up of the underlying AIIRD occurred within two months after vaccination in all the other patients.

**Figure 1:**
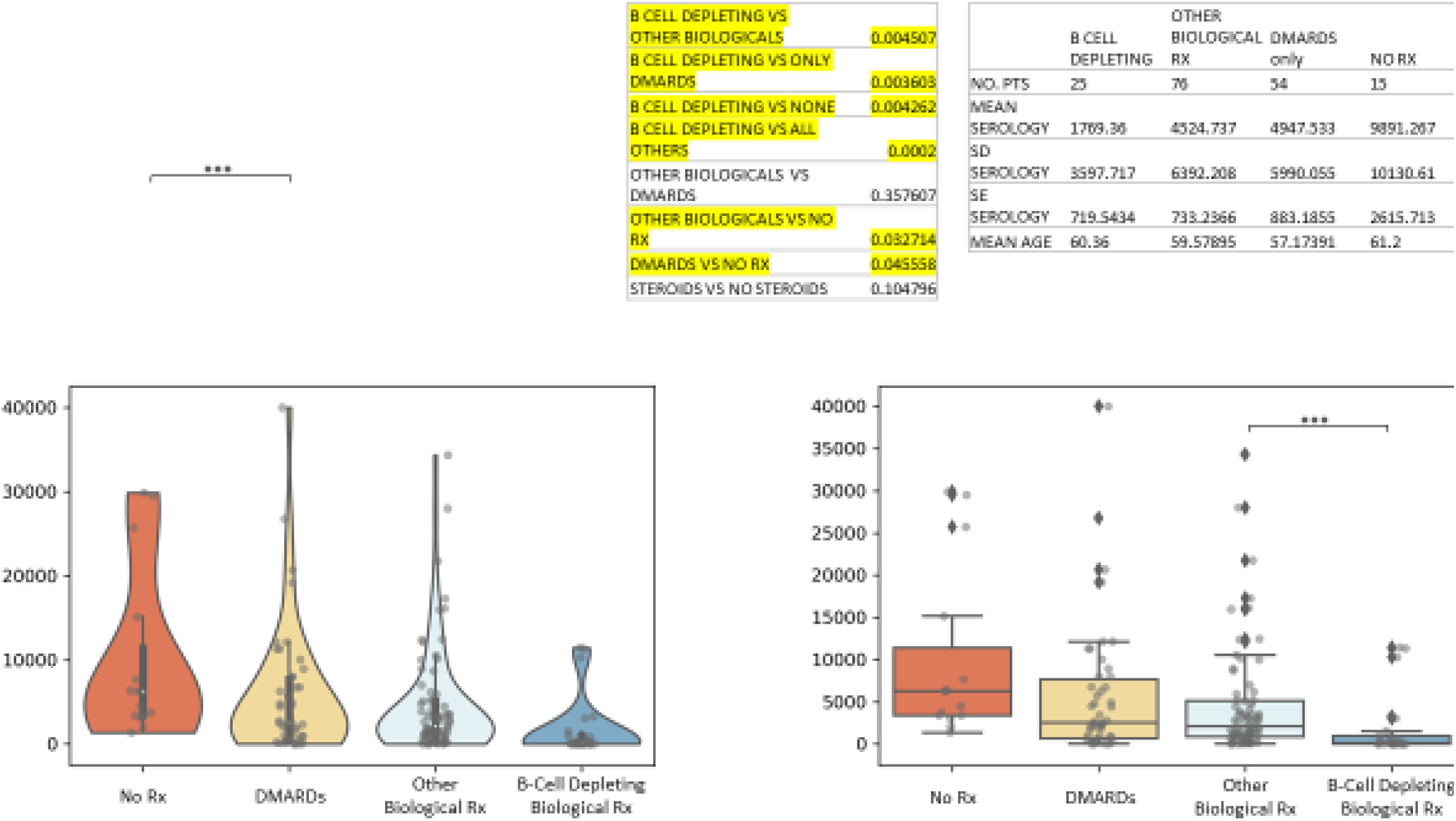
Serology per drug (vaccinated) Legend: Antibody titers for the different treatments presented as boxplots, indicating the median and quartiles with whiskers up to 1.5 times the interquartile range. The violin plot outlines illustrate kernel probability density/the distribution of the data.

**Figure 2:**
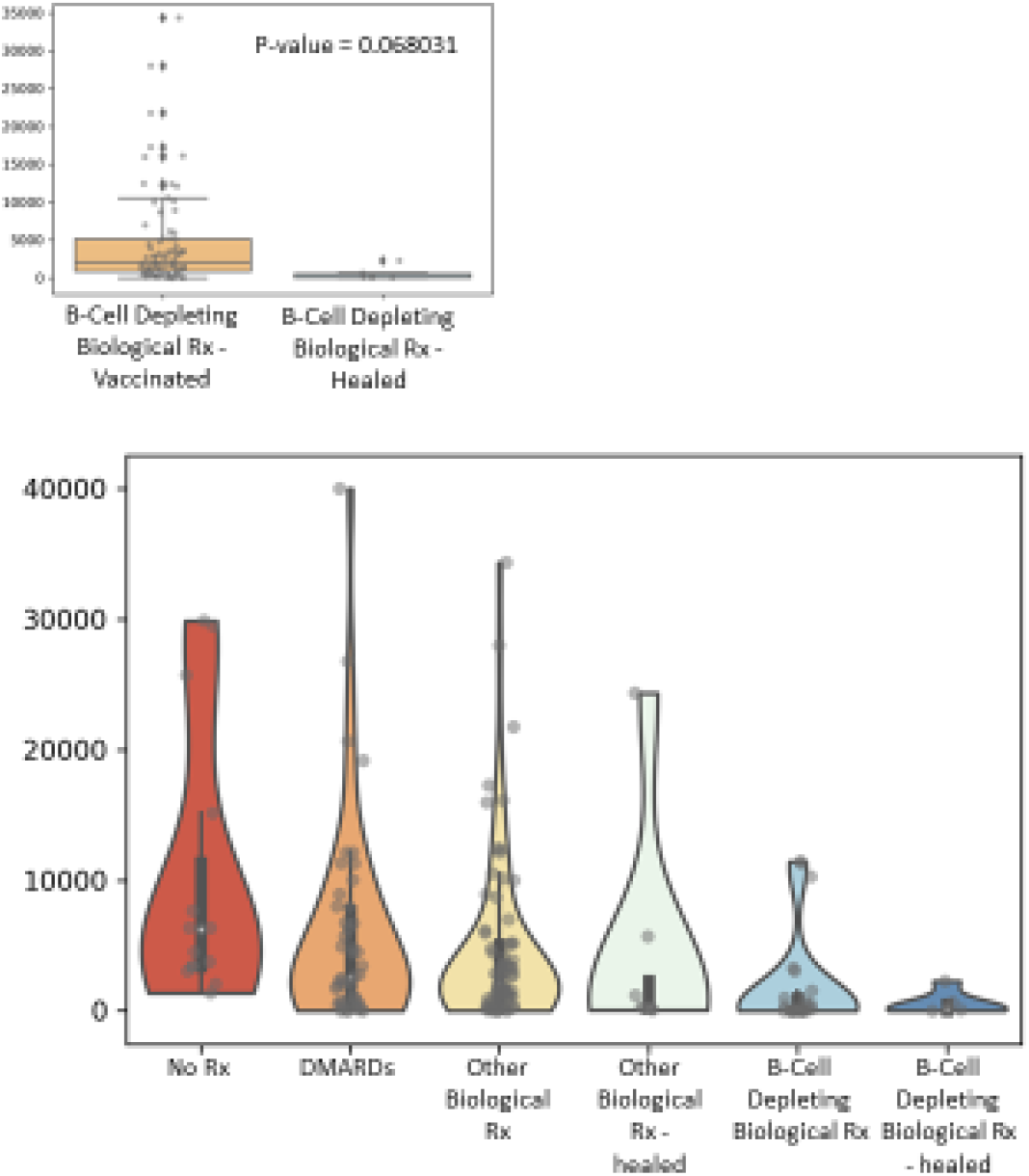
Serology per drug (vaccinated + healed patients)

The Pfizer mRNA vaccine against SARS CoV2 virus appears to be safe in our patients, only minor side effects were reported and no apparent impact on AIIRD activity was noted. The vast majority of AIRD patients developed a significant humoral response following the administration of the second dose of the vaccine. The antibody levels were influenced by the medication used to treat the AIIRD and not by the type or activity of the AIIRD. Untreated and DMARDs treated patients mounted similar antibody titers that were about one log higher than the patients treated with biologics and MMF. Notably, 68% of the non-responders were treated with B cell depleting agents or MMF. Further studies should assess whether the low antibody titers are associated with diminished protection against COVID19 severe disease and whether the timing of anti CD20 Ab administration has an effect upon the anti S Ab titer.

## Data Availability

Contact the lead author for data.

